# Disability-adjusted life years (DALYs) due to the direct health impact of COVID-19 in India, 2020

**DOI:** 10.1101/2021.08.20.21262326

**Authors:** Balbir B Singh, Brecht Devleesschauwer, Mehar S Khatkar, Mark Lowerison, Baljit Singh, Navneet K Dhand, Herman W Barkema

**Affiliations:** Centre for One Health, Guru Angad Dev Veterinary and Animal Sciences University, Ludhiana, Punjab 141004, India; Sydney School of Veterinary Science, The University of Sydney, 425 Werombi Road, Camden, 2570 NSW, Australia; Dept. of Veterinary Microbiology, University of Saskatchewan, Saskatoon, Canada; Department of Epidemiology and Public Health, Sciensano, Rue Juliette Wytsman 14, BE-1050, Brussels, Belgium; Department of Veterinary Public Health and Food Safety, Ghent University, Salisburylaan 133, BE-9820, Merelbeke, Belgium; Dept. of Community Health Sciences, Cumming School of Medicine, University of Calgary, 3330 Hospital Drive NW, Calgary, AB, Canada, T2N 4N1; One Health at UCalgary, University of Calgary, 3330 Hospital Drive NW, Calgary, Canada, T2N 4N1; University of Saskatchewan, Saskatoon, SK, Canada; Dept. of Production Animal Health, Faculty of Veterinary Medicine, University of Calgary, 3330 Hospital Drive NW, Calgary, Canada, T2N 4N1

**Keywords:** COVID-19, Disability-adjusted life years, India, Years lived with disability, Years of life lost

## Abstract

COVID-19 has affected all countries. Its containment represents a unique challenge for India due to a large population (>1.38 billion) across a wide range of population densities. Assessment of the COVID-19 disease burden is required to put the disease impact into context and support future pandemic policy development. Here, we present the national-level burden of COVID-19 in India in 2020 that accounts for differences across urban and rural regions and across age groups. Disability-adjusted life years (DALY) due to COVID-19 were estimated in the Indian population in 2020, comprised of years of life lost (YLL) and years lived with disability (YLD). Scenario analyses were conducted to account for excess deaths not recorded in the official data and for reported COVID-19 deaths. The direct impact of COVID-19 in 2020 in India was responsible for 14,106,060 (95% uncertainty interval [UI] 14,030,129–14,213,231) DALYs, consisting of 99.2% (95% UI 98.47–99.64%) YLLs and 0.80% (95% UI 0.36–1.53) YLDs. DALYs were higher in urban (56%; 95% UI 56–57%) than rural areas (44%; 95% UI 43.4–43.6) and in males (64%) than females (36%). In absolute terms, the highest DALYs occurred in the 51–60-year-old age group (28%) but the highest DALYs per 100,000 persons were estimated for the 71-80 year old age group (5,481; 95% UI 5,464–5,500 years). There were 4,823,791 (95% UI 4,760,908–4,924,307) DALYs after considering reported COVID-19 deaths only. The DALY estimations have direct and immediate implications not only for public policy in India, but also internationally given that India represents one sixth of the world’s population.

## 1.0 Introduction

The coronavirus disease 2019 (COVID-19) pandemic ^1^ originated in Wuhan, China in December 2019 ^2^ and eventually worldwide. The disease is caused by the severe acute respiratory syndrome coronavirus 2 (SARS-CoV-2) virus that infects humans ^3^. As of 30 July 2021, 196,553,009 cumulative cases and 4,200,412 cumulative deaths have been reported worldwide ^4^. The virus spreads via respiratory droplets, and aerosols shed from infected individuals ^5,6^ or directly through contaminated hands ^7^. Important symptoms include fever, respiratory tract symptoms such as cough, pneumonia, shortness of breath and can progress to serious illness and even death ^8,9^. In addition, long or post-acute COVID-19 syndrome with symptoms and abnormalities after 12 weeks following the onset of acute COVID-19 infections has also been recognised ^10^.

The first case of COVID-19 in India was reported on 30 January 2020. The government of India implemented staged lockdowns and intense contact mitigation to overcome this challenge; however, these interventions were only able to slow but did not halt the spread of COVID-19 in India ^11^. As of 30 July 2021, 31,572,344 cumulative cases and 423,217 cumulative deaths have been reported in India ^4^. The adjusted case fatality risk (aCFR) due to COVID-19 in India has been reported to be 1.4 and 3.0% using random- and fixed-effects models, respectively ^12^. At the end of the first wave (15 February 2021), the cumulative aCFR declined to 1.2% using random and 1.6% using fixed-effects models ^12^. The aCFR in India was associated with co-morbidities such as the incidence of diabetes, cardiovascular diseases, hypertension, and acute respiratory infections ^12^.

The health impact of COVID-19 is being investigated across the globe. A recent study reported 1,279,866 deaths due to COVID-19 in 81 countries with an average of 16 years of life lost (YLL) per death resulting in 20,507,518 YLL ^13^. In the Kerala state of India, 709.2 DALYs per 100,000 population have been estimated from the inception of the pandemic to 20 August 2020 ^14^. However, despite being the second most populous country in the world, no comprehensive assessment of the disease burden of COVID-19 in India has been undertaken. In this study, we estimated DALYs due to the direct health impact of COVID-19 in India from 30 January 2020 (the first case reported) to 31 December 2020. Domain, gender, and age-specific DALY estimates are provided to understand health impacts of COVID-19 in India.

## 2.0 Methods

### 2.1 COVID-19 case and death data

The COVID-19 cumulative case and death data till 31 December 2020 were extracted using the Government of India Ministry of Health and Family Welfare (MoHFW) COVID-19 tracker on 18 January 2021, 8:00 AM ^15^. Age-, gender- and domain-specific distributions of COVID-19 cases as provided by the National Centre for Disease Control, Government of India (as of 18 January 2021) were used in the analysis ^16^. The National Centre for Disease Control data on the proportion of deaths in various age groups as of 2 September 2020 were recorded from the published literature ^17^. Gender-specific mortality risk (64% in males and 36% in females) reported by the MoHFW were also recorded as of 21 May 2020 ^18^. Domain-specific COVID-19 death data (rural vs urban) were obtained from the news item published online (https://www.downtoearth.org.in/news/health/more-than-half-of-india-s-april-covid-19-deaths-were-in-rural-districts-76782) indicating 56% deaths in urban and 44% deaths in rural districts during March 2020-April 2021.

Several studies indicate 48.5% ^19^, 52.5% ^20^, and 57.8% ^21^ of SARS-CoV-2 infected individuals remained asymptomatic in the country. A systematic review and meta-analysis conducted using the studies conducted worldwide till 10 June 2020 indicated 31% (95% CI 26–37%) of infected people to be asymptomatic ^22^. Based on all these studies, we assumed that 31–57.8% of infected people to be asymptomatic in the country.

As of 17 July 2020, the MoHFW reported that there were 2.8% cases on oxygen beds, 1.9% cases in intensive care units (ICUs), and 0.35% cases on ventilators ^23^. As of September 4, 2020, the MoHFW reported updated estimates of 3.5% cases on oxygen beds, 2.0% cases in ICUs, and 0.50% cases on ventilators ^23^. Based on both estimates, we assumed 4.8-5.5% of severe (patients in ICU plus those needing oxygen support), and 0.35-0.5% of critical (patients on ventilators) cases in the country. The proportion of cases that were not asymptomatic, severe or critical were assumed to be mild/moderate cases.

No data were available for the proportion of cases having post-acute COVID-19 syndrome (PCS) from India. A UK COVID symptom study indicates that approximately 10% of COVID-19 patients suffer from PCS ^24,25^. Other studies report that 51% of the patients in Spain ^26^ and 32.5% of patients in the United states ^27^ suffer from PCS. Based on the above-mentioned findings, we assumed that 32.5% of the mild/moderate, severe or critical cases would suffer from PCS in India. Details of COVID-19 cases and death input data have been presented in Table S1.

### 2.2 COVID-19 severity data

COVID-19 outcomes model and value of health loss as described by Wyper, et al. ^28^ were used in the current estimations ^29,30^. For COVID-19 infections, disability weight(s) of 0.051 (0.032-0.074) for mild/moderate, 0.133 (0.088-0.190) for severe, 0.655 (0.579-0.727) for critical, and 0.219 (0.148-0.308) for post-acute consequences cases were used. Per definition, the disability weight for asymptomatic cases equaled zero.

A mean duration of 7.79 days (6.20 - 9.64 days) has been estimated using meta-analysis for lower respiratory infections (LRI) ^31^. A study from India reported the interval to resolution of COVID-19 symptoms to be a median of 7.0 days (2.0-9.5 days) ^32^. Similarly, another study reported a median moderate COVID-19 duration of 6.0 days (3-11 days) ^33^. Based on all these estimates, we used a median of 7 days (2-11 days) as the disease duration for mild/moderate COVID-19 in India.

A study from India reported 12 days duration of COVID-19 for hospitalized patients ^34^. Another study from Germany also reported an overall hospitalization of 12 (7–20) days for COVID-19 patients needing oxygen supplementation outside the ICU ^35^. For critical patients, duration in the ICU for 9 (7-11) days has been reported ^36^. A Belgian study reported the time between symptom onset and hospitalization to vary from 3-10.4 days ^37^. Therefore, we used 18 (10-30.4) days as the disease duration for severe and critical COVID-19 (symptom onset to hospitalization duration plus hospitalization duration) in India.

A recent review on PCS indicated the duration of PCS in various countries varied from 1 to 6 months post-symptom onset, with a 2-month duration commonly reported ^10^. Therefore, we used a median of 60 days (30 – 180 days) as the disease duration for PCS in India. Details of COVID-19 severity-related input data have been presented in Table S1.

### 2.3 Demographic data

The human population in India for 2020 was projected to be 1,380,004,390 ^38^. However, age-, domain- and gender-specific proportions of the population were not available for the year 2020. Therefore, we used the age-, domain- and gender-specific proportion of the population from the 2011 human population census data ^39^ assuming it to be constant for 2011 and 2020 population data. Life expectancy was derived as per the standard life expectancy table of Global Burden of Disease Study 2019 ^40^. Details of demographic data have been presented in Table S1.

### 2.4 COVID-19 deaths

Excess deaths due to COVID-19 in India for the year 2020 were estimated (Table S1) using COVID-19 mortality estimates from March 2020 to May 2021 generated by the Institute for Health Metrics and Evaluation, Washington ^41^. Using the IHME data, the confidence interval of the proportion of the reported COVID-19 deaths to the excess deaths was estimated ^42^. We assumed that there will be a similar proportion of excess deaths during the year 2020 as reported from March 2020 to May 2021. As age, domain, and gender-specific proportion of excess deaths were unavailable, we assumed that the proportion of excess deaths will be similar to those reported for COVID-19 deaths.

### 2.5 Calculation of disability-adjusted life years

DALYs were estimated as the sum of YLDs and YLLs. YLL was estimated by multiplying the number of deaths due to COVID-19 by the residual standard life expectancy at the age of death due to the disease. YLD was calculated as a product of the number of incident cases of COVID-19, disease duration and disability weight. Domain-, gender- and age-specific DALYs were estimated as per Devleesschauwer et al. 2014a and 2014b ^43,44^. We assumed that cases were sick before dying; therefore, both YLDs and YLLs were estimated for COVID-19 deaths.

Scenario analyses were conducted using a) excess COVID-19 deaths reported by IHME ^41^, and b) officially reported deaths ^15^ to estimate DALYs associated with COVID-19 in India. Uncertainty was propagated using 10,000 Monte Carlo simulations. Uniform probability distributions were applied for the proportion of patients having mild/moderate, severe, or critical symptoms. Triangular distribution(s) were applied to disability weights and COVID-19 disease duration associated with various COVID-19 outcomes. A triangular distribution was also applied to excess COVID-19 deaths during the calendar year 2020.

The resulting uncertainty distributions were summarized by their mean and a 95% uncertainty interval (UI) defined as the distribution’s 2.5^th^ and 97.5^th^ percentile.

All analyses were conducted using R version 3.6.3 (R Development Core Team, http://www.r-project.org).

## 3.0 Results

### 3.1 Disability-adjusted life years

The direct impact of COVID-19 was responsible for 14,106,060 DALYs consisting of 13,992,803 (99.2%; 95% UI 98.47–99.64%) YLLs and 113,257 (0.80%; 95% UI 0.36– 1.53%) YLDs in 2020 in India (Table 1). There were 7,970,337 DALYs in urban (56%) as compared to 6,135,722 in rural (44%) areas (Table 1). There were 9,028,492 DALYs in males (64.0%) as compared to 5,077,568 DALYs in females (36.0%) (Table 1). The highest DALYs were reported in persons 51-60 years (3,922,508; 28%, Table 2). Mortality contributed towards almost all (99.2%) DALYs due to the direct impact of COVID-19 in India. There were 31.67 YLLs per COVID-19 death in the country. PCS contributed towards 89.61% (95% UI 79.1–95.62%) of the YLDs.

**Table 1:** Estimated annual disability adjusted life years (DALYs) due to excess COVID-19 deaths in 2020 in India.

|  | **Median YLL** | **95% UI** | **Median YLD** | **95% UI** | **Median DALY** | **95% UI** |
| --- | --- | --- | --- | --- | --- | --- |
| **Total** | 13,992,709 | 13,958,073–<br>14,027,485 | 105,784 | 50,002–217,970 | 14,100,422 | 14,030,129–<br>14,213,231 |
| **Domain** |  |  |  |  |  |  |
| *Rural* | 6,102,220 | 6,087,116–<br>6,117,386 | 31,253 | 14,773–64,397 | 6,134,167 | 6,109,995–<br>6,168,716 |
| *Urban* | 7,890,488 | 7,870,957–<br>7,910,099 | 74,530 | 35,229–153,573 | 7,966,143 | 7,919,750–<br>8,045,000 |
| **Gender** |  |  |  |  |  |  |
| *Male* | 8,955,334 | 8,933,167–<br>8,977,590 | 68,275 | 32,273–140,681 | 9,024,833 | 8,979,611–<br>9,097,629 |
| *Female* | 5,037,375 | 5,024,906–<br>5,049,895 | 37,508 | 17,729–77,289 | 5,075,566 | 5,050,493–<br>5,115,612 |
|  | **Median**<br>**YLL/100,000**<br>**person years** | **95% UI** | **Median**<br>**YLD/100,000**<br>**person years** | **95% UI** | **Median**<br>**DALY/100,000**<br>**person years** | **95% UI** |
| **Total** | 1,014 | 1,011–1,016 | 8 | 4–16 | 1,022 | 1,017–1,030 |
| **Domain** |  |  |  |  |  |  |
| *Rural* | 642 | 641–644 | 3 | 2–7 | 646 | 643–649 |
| *Urban* | 1,836 | 1,831–1,840 | 17 | 8–36 | 1,854 | 1,842–1,872 |
| **Gender** |  |  |  |  |  |  |
| *Male* | 1,261 | 1,258–1,264 | 10 | 5–20 | 1,270 | 1,264–1,281 |
| *Female* | 752 | 750–754 | 6 | 3–12 | 758 | 754–764 |

**Table 2:** Estimated annual disability-adjusted life years (DALYs) due to excess COVID-19 deaths in various age groups in 2020 in India.

| Age (years) | Median<br>DALYs | 95% UI | Median DALY<br>per 100,000<br>person-years | 95% UI |
| --- | --- | --- | --- | --- |
| **Total** | 14,100,422 | 14,030,129–<br>14,213,231 | 1022 | 1,017–1,030 |
| *0-10* | 189,636 | 187,389–194,019 | 61 | 61–63 |
| *11-20* | 237,600 | 233,106–246,611 | 83 | 81–86 |
| *21-30* | 760,728 | 748,089–785,869 | 311 | 306–322 |
| *31-40* | 1,485,568 | 1,472,653–1,509,745 | 758.4 | 752–771 |
| *41-50* | 2,652,051 | 2,639,491–2,671,686 | 1,822 | 1,814–1,836 |
| *51-60* | 3,921,906 | 3,908,098–3,939,808 | 4,069 | 4,055–4,088 |
| *61-70* | 3,265,799 | 3,255,655–3,277,308 | 5,001 | 4,986–5,019 |
| *71-80* | 1,326,741 | 1,322,679–1,331,288 | 5,481 | 5,464–5,499 |
| *81-90* | 245,109 | 244,293–246,096 | 3,805 | 3,793–3,821 |
| *> 90 years* | 15,072 | 14,996–15,194 | 714 | 710–720 |

95% UI = 2.5–97.5th percentile

### 3.2 Disability-adjusted life years per 100,000 people

There were 1,022 DALYs, 1,014 YLLs, and 8 median YLDs per 100,000 persons per year in the country (Table 1). The median DALYs per 100,000 persons per year were higher in urban as compared to rural (1,854 *vs* 646) and in males as compared to females (1,270 vs 758) in India. The highest DALYs per 100,000 persons per year were at 5,481 (95% UI 5,464–5,500) reported in persons belonging to the age group of 71-80 years in India (Table 2).

### 3.3 Scenario analyses

The DALYs, YLLs and YLDs were approximately 3 times lower after considering reported COVID-19 deaths only (Table S2). There were 4,823,791 DALYs, 4,710,106 YLLs and 113,685 YLDs due to reported COVID-19 deaths. There were 349 DALYs, 341 YLLs and 8 YLDs per 100,000 persons per year due to reported COVID-19 deaths in the country (Table S2). A proportional decrease in age-wise DALYs and YLLs after excluding excess COVID-19 deaths is also presented in Table S3.

## 4.0 Discussion

We estimated DALYs due to the direct impact of COVID–19 in 2020 in India. Furthermore, we also accounted for excess COVID-19 deaths (not officially reported) for accurate DALY estimates. As far as we are aware, DALY estimations for COVID-19 for the full 2020 year have not been reported from India. Therefore, the current study will provide valuable insights into health impacts of COVID-19 in 2020 in India.

We believe IHME excess COVID-19 deaths prediction to be accurate, as it was based on models using week-by-week measurements of excess death rate during the ongoing COVID-19 pandemic. Furthermore, 6 drivers of all-cause mortality, including: a) the excess COVID-19 death rate, b) excess mortality due to delayed or deferred health care, c) excess mortality due to mental health disorders, d) reductions in mortality due to decrease in injuries, e) decrease in mortality due to other viruses, and f) and decrease in mortality due to certain chronic conditions, were also taken into account to accurately predict excess COVID-19 deaths nationally and various regions globally ^41^.

There was a loss of 1,022 healthy years of life per 100,000 person-years and 14 million DALYs due to the direct impact of COVID-19 in 2020 in India. In 2019, there were overall 468 million DALYs in India ^45^ indicating that the DALYs due to COVID-19 could account for almost 3% of the total DALYs in India. In 2017, the top 15 causes of DALYs per 1,000 person-years in India ranges from ischaemic heart disease (35.0) to fever of unknown origin (9.0) ^46^. Therefore, if excess deaths due to COVID-19 were accounted, the direct impact of COVID-19 would be among the top 15 causes but not among the top 10 of DALYs in 2020 in India. These estimates of direct impact of COVID-19 are lower than ischaemic heart disease, nutritional deficiencies, chronic respiratory diseases, neuropsychiatric conditions, diarrhoea, vision and other sensory loss, respiratory infections, cancers, stroke, road traffic accidents, tuberculosis, liver and alcohol-related conditions, but higher than musculoskeletal disorders and fever of unknown origin.

There are considerable variations among countries in estimated DALYs. For example, there were 1,767–1,981 DALYs per 100,000 person-years due to COVID-19 during 2020 in Scotland ^47^, 4.9 (20 January to 24 April 2020) in Korea ^48^, and 368 in Germany in 2020 ^49^. There were 1200 YLLs per 100,000 capita during the first year of the pandemic in USA ^50^. Based on the reported COVID-19 death data, the DALYs per 100,000 person-years were approximately similar to those reported for Germany. However, after accounting for excess COVID-19 deaths, the DALYs per 100,000 person-years were much closer to those reported from the USA.

Median DALYs were higher in urban areas (56%) compared to rural areas. Similarly, urban-rural differences in exposures and outcomes of COVID-19 have been reported in the USA ^51^. This is because a high number of cases and deaths were reported from urban as compared to rural areas. Perhaps contact patterns are different in urban areas and rural areas. For example, a social contact rate of 17.0 in rural ^52^, 28.3 in urban developed centres and 67.4 in urban slum areas ^53^ per person per day was recorded in various domains in India.

Median DALYs were higher in males (64.0%) as compared to females (36.0%). Similar trends have been observed globally. Out of 121,449 DALYs in Italy, 82,020 were in males and 39,429 in females ^54^. Higher YLLs have been reported in males in 30 high incidence countries ^55^. Gender-specific DALY differences are due to a difference in proportion in cases and deaths among males and females. Furthermore, women have fewer contacts than men outside the home ^52^. In addition, being a male has been identified as a risk factor for death and ICU admission due to COVID-19 ^56^.

Mortality contributed (99.61%) towards almost all the DALYs due to the direct impact of COVID-19 in India, in line with other studies ^49,54,57^. There was a median of 31.67 YLLs per COVID-19 death in India. This is because highest median YLLs were estimated for the population in the 51-60 year (3,906,065) and 61-70 year (3,256,690) age-groups, respectively. A study from two Indian states also reported that cases and deaths are concentrated in younger cohorts in the country as compared to high-income countries ^58^. Our DALY estimations were derived using the standard life expectancy table of Global Burden of Disease Study 2019 ^40^. In Germany, 9.6 YLL per death have been reported; however, a higher YLL per death of 25.2 have been reported for persons under 70 years of age ^49^. YLL per COVID-19 death ranged from 15.3 to 15.5 in Scotland ^59^. However, these estimates could not be compared due to different life expectancy tables used in the analysis.

There are uncertainties surrounding these estimates. Undercounting or underreporting of COVID-19 cases and deaths is a serious concern and is more relevant in low- and middle-income countries such as India ^60^. We used IHME excess COVID-19 deaths and officially reported data to drive our estimates. As exact data on the total number of deaths during 2020 remain unavailable (India’s Civil Registration System data), DALYs after accounting for the exact number of excess deaths could not be estimated. Grey literature indicates that there were 13,297 more deaths during March-September 2020 as compared to the same period in 2019 and 14,944 as compared in 2018 in Mumbai, India (https://timesofindia.indiatimes.com/city/mumbai/13k-more-deaths-in-city-this-year-between-march-sept/articleshow/78920631.cms). Conversely, the Kerala state of India reported an 11.1% reduction in all-cause mortality during 2020 as compared to 2019 ^61^ possibly due to high reduction in other cause mortality as compared to COVID-19 associated excess mortality. Globally, comparative analyses using COVID-19 attributable and excess death approaches indicate that YLL estimations using attributable deaths maybe 3 times lower than those accounting for excess deaths; however, these estimates maybe even 12 times lower in some countries ^13^. To overcome this issue, we used excess mortality due to COVID-19 from March 2020 to May 2021 as estimated by the IHME, Washington ^41^. In addition, no data were available on age- and domain-specific proportions of excess deaths. We assumed this proportion to be similar to reported deaths. Therefore, the availability of all-cause mortality data in future will further refine COVID-19 associated DALY estimations in India. Furthermore, no data were available on the proportion of post-acute covid manifestations and was derived from other studies. This might have resulted in over or under estimation in the presented DALY. Lastly, indirect impact of COVID-19 especially due to disruption of health services could not be estimated.

Overall, direct impact of COVID-19 could be among the top 15 causes of DALYs in India. However, the estimated impact of COVID-19 would be substantially higher if indirect impacts of COVID-19 were to be accounted. Health impacts of COVID-19 varies in substantially in urban and rural areas. The current study will help to understand health impacts of COVID-19 in India and in other regions and countries.

## Data Availability

The analysed data are available along with the manuscript. Sources of the raw data used in the analysis have been cited.

## ACKNOWLEDGEMENTS

The authors acknowledge India’s National and State Health departments for collecting daily COVID-19 epidemic data and releasing it in the public domain.

## ETHICAL STATEMENT

Informed consent for collection of epidemiological data was not required, as these data were already coded and available in the public domain. No identifiable personal information was used in this study.

## AUTHOR CONTRIBUTORS

BBS conceived the study. NKD, MSK and BD contributed to the conceptual model development. BBS build the model and conducted the analyses. BBS prepared the first draft of the paper which was reviewed and edited by NKD, MSK, HWB, ML, BS and BD. HWB and NKD coordinated the project and preparation of the manuscript.

