## Supplementary material for "Disability-adjusted life years (DALYs) due to the direct health impact of COVID-19 in India, 2020": Table S1

**Supplementary appendix**

**Table S1. Input parameters used to estimate disability adjusted life years due to COVID-19 in India in 2020.**

| **Parameter** | **Value** | **Distribution** | **Reference** |
| --- | --- | --- | --- |
| ***Population parameters*** |  |  |  |
| Human population in India, 2020 | 1,380,004,390 | Fixed | World Bank 2020 |
| *Age wise distribution of human population in India (years)* |  |  |  |
| ≤ 10 | 22.32% | Fixed | COI, 2011 |
| 11-20 | 20.77% | Fixed | COI, 2011 |
| 21-30 | 17.70% | Fixed | COI, 2011 |
| 31-40 | 14.19% | Fixed | COI, 2011 |
| 41-50 | 10.54% | Fixed | COI, 2011 |
| 51-60 | 6.98% | Fixed | COI, 2011 |
| 61-70 | 4.73% | Fixed | COI, 2011 |
| 71-80 | 1.75% | Fixed | COI, 2011 |
| 81-90 | 0.47% | Fixed | COI, 2011 |
| ≥ 90 | 0.15% | Fixed | COI, 2011 |
| *Domain and gender wise distribution of human population in India* |  |  |  |
| Rural, total | 68.86% | Fixed | COI, 2011 |
| Rural, male | 35.33% | Fixed | COI, 2011 |
| Rural, female | 33.53% | Fixed | COI, 2011 |
| Urban, total | 31.14% | Fixed | COI, 2011 |
| Urban, male | 16.14% | Fixed | COI, 2011 |
| Urban, female | 15.0% | Fixed | COI, 2011 |
| **COVID-19 parameters** |  |  |  |
| Cumulative cases and deaths (officially reported) | 10,266,674 and 148,738 | Fixed | MIB, 2020 |
| Excess COVID-19 deaths (March 2020 to May 2021) | 736,811 | Fixed | IHME, 2021 |
| Reported COVID-19 deaths (March 2020 to May 2021) | 248,016 | Fixed | IHME, 2021 |
| Excess COVID-19 deaths (calendar year 2020) | 441,874 (440,462-443,295 | Triangular | calculation – see text |
| *Age wise distribution of COVID-19 cases and deaths in India** |  |  | NCDC, 2021; Indrayan et al., 2020 |
| ≤ 10 years | 3.9% and 0.5% | Fixed |  |
| 11-20 years | 7.99% and 0.7% | Fixed |  |
| 21-30 years | 22.33% and 2.6% | Fixed |  |
| 31-40 years | 21.63% and 6.1% | Fixed |  |
| 41-50 years | 17.33% and 13.4% | Fixed |  |
| 51-60 years | 14.52% and 25.3% | Fixed |  |
| 61-70 years | 8.24% and 28.6% | Fixed |  |
| 71-80 years | 3.19% and 17% | Fixed |  |
| 81-90 years | 0.77% and 5.3% | Fixed |  |
| ≥ 90 years | 0.09% and 0.5% | Fixed |  |
| *Gender wise distribution of COVID-19 cases and deaths in India** |  |  | NCDC, 2021; PIB, 2020b |
| Male | 64.55% and 64% | Fixed |  |
| Female | 35.45% and 36% | Fixed |  |
| *Domain wise distribution of COVID-19 cases and deaths in India** |  |  | NCDC, 2021; see text |
| Rural | 29.55% and 44% | Fixed |  |
| Urban | 70.45% and 56% | Fixed |  |
| ***Disease severity parameters*** |  |  |  |
| *Proportion of COVID-19 cases* |  |  | See text |
| Asymptomatic | 31%-57.8% | Uniform |  |
| Mild/moderate | [100-(asymptomatic + severe + critical)]% | Uniform |  |
| Severe | 4.8%-5.5% | Uniform |  |
| Critical | 0.35%-0.5% | Uniform |  |
| Post-acute Covid consequences | 32.5% of the mild/moderate, severe or critical cases | Uniform | Calculation; See text |
| *Disability Weight* |  |  | Wyper et al., 2021c; Haagsma et al., 2015; Salomon et al., 2015 |
| Asymptomatic | - | - | - |
| Mild/moderate | 0.051 (0.032–0.074) | Triangular |  |
| Severe | 0.133 (0.088–0.19) | Triangular |  |
| Critical | 0.655 (0.579–0.727) | Triangular |  |
| Post-acute Covid consequences | 0.219 (0.148–0.308) | Triangular |  |
| *Duration of disability* |  |  | See text |
| Asymptomatic | - | - | - |
| Mild/moderate | 7 (2-11) days | Triangular |  |
| Severe | 18 (10-30.4) days | Triangular |  |
| Critical | 18 (10-30.4) days | Triangular |  |
| Post-acute Covid consequences | 60 (30 – 180 days) | Triangular |  |
| *Life expectancy (approximated)* |  |  | GBDCN, 2021 |
| ≤ 10 years | 83.96 | Fixed |  |
| 11-20 years | 74.08 | Fixed |  |
| 21-30 years | 64.155 | Fixed |  |
| 31-40 years | 54.26 | Fixed |  |
| 41-50 years | 44.475 | Fixed |  |
| 51-60 years | 34.94 | Fixed |  |
| 61-70 years | 25.77 | Fixed |  |
| 71-80 years | 17.615 | Fixed |  |
| 81-90 years | 10.43 | Fixed |  |
| ≥ 90 years | 6.77 | Fixed |  |

*Cases not specified proportionally divided

**Table S2: Estimated annual disability adjusted life years (DALY) due to the reported COVID-19 deaths in 2020 in India.**

|  | **Median YLL** | **95% UI** | **Median YLD** | **95% UI** | **Median DALY** | **95% UI** |
| --- | --- | --- | --- | --- | --- | --- |
| **Total** | 4,710,106 | 4,710,106–4,710,106 | 105,803 | 50,803–214,201 | 4,815,908 | 4,760,908–4,924,307 |
| **Domain** | | |  |  |  |  |
| *Rural* | 2,054,077 | 2,054,077–2,054,077 | 31,259 | 15,009–63,284 | 2,085,336 | 2,069,087–2,117,361 |
| *Urban* | 2,656,029 | 2,656,029–2,656,029 | 74,544 | 35,793–150,917 | 2,730,573 | 2,691,822–2,806,946 |
| **Gender** | | |  |  |  |  |
| *Male* | 3,014,468 | 3,014,468–3,014,468 | 68,288 | 32,789–138,248 | 3,082,755 | 3,047,257–3,152,716 |
| *Female* | 1,695,638 | 1,695,638–1,695,638 | 37,515 | 18,013–75,952 | 1,733,153 | 1,713,651–1,771,590 |
|  | **Median YLL/100,000 person years** | **95% UI** | **Median YLD/100,000 person years** | **95% UI** | **Median DALY/100,000 person years** | **95% UI** |
| **Total** | 341 | 341–341 | 8 | 4–16 | 349 | 345–357 |
| **Domain** |  |  |  |  |  |  |
| *Rural* | 216 | 216– 216 | 3 | 2–7 | 219 | 218–223 |
| *Urban* | 618 | 618– 618 | 17 | 8–35 | 635 | 626–653 |
| **Gender** | | |  |  |  |  |
| *Male* | 424 | 424– 424 | 10 | 5–19 | 434 | 429–444 |
| *Female* | 253 | 253– 253 | 6 | 3–11 | 259 | 256–265 |

95% UI = 2.5–97.5^th^ percentile

**Table S3: Estimated annual disability-adjusted life years (DALYs) due reported COVID-19 deaths in different age groups in 2020 in India.**

| **Age (in years)** | **Median DALYs** | **95% UI** | **Median DALY per 100,000 person years** | **95% UI** |
| --- | --- | --- | --- | --- |
| **Total** | 4,815,908 | 4,760,908–4,924,307 | 349 | 345–357 |
| *0-10* | 66,566 | 64,421–70,793 | 22 | 21–23 |
| *11-20* | 85,583 | 81,188–94,243 | 30 | 28–33 |
| *21-30* | 271,724 | 259,443–295,927 | 111 | 106–121 |
| *31-40* | 515,186 | 503,290–538,630 | 263 | 257–275 |
| *41-50* | 904,761 | 895,230–923,545 | 622 | 615–635 |
| *51-60* | 1,330,179 | 1,322,193–1,345,917 | 1380 | 1372–1397 |
| *61-70* | 1,104,949 | 1,100,418–1,113,881 | 1692 | 1685–1706 |
| *71-80* | 448,778 | 447,023–452,235 | 1854 | 1847–1868 |
| *81-90* | 83,036 | 82,612–83,870 | 1289 | 1283–1302 |
| *> 90 years* | 5,148 | 5,088–5,265 | 244 | 241–249 |

95% UI = 2.5–97.5th percentile
